# Evaluating large language models for natural-language-to-code generation on aggregate Czech public health data analysis

**DOI:** 10.64898/2025.12.05.25341697

**Authors:** Ondrej Klempir, Ladislav Dusek, Radim Krupicka, Gleb Donin, Jan Zigmond, Radka Storchova, Ales Tichopad

## Abstract

Large language models (LLMs) are increasingly explored as tools for healthcare research and data analysis. However, their applicability to structured public health datasets, especially in non-English contexts, remains underexamined. We systematically evaluated 11 state-of-the-art LLMs on their ability to generate executable Python code for analytical queries over Czech public health datasets, focusing on incidence and prevalence data provided by the National Health Information Portal (known as NZIP). A set of representative analytical queries were designed, covering filtering, aggregation, weighted averages, and identification of primary diagnoses. Each model was prompted in Czech and assessed on code executability, correctness of results, and ability to adapt to local terminology. In the majority of cases, the models generated syntactically valid code within one minute, but performance varied. For the main objective of replicating “ground truth” queries as per dataset documentation, ChatGPT-4o achieved the highest accuracy, followed closely by GPT-4.1 mini. Claude and Gemini models frequently failed to apply critical filtering instructions, while Deepseek-R1, though accurate, defaulted to English output. Some models produced code that executed successfully but returned incorrect results, underscoring the need for systematic validation. Overall, LLMs show strong potential as coding assistants in public health analytics, even in Czech-language settings. Their integration into hybrid human–AI workflows, combined with validation mechanisms and retrieval-augmented generation, may accelerate the creation of reliable analytical pipelines.

## 1. Introduction

Large language models (LLMs) are increasingly permeating key tasks in biomedical natural language processing (NLP) applications [1]. Recently, an article introduced Delphi-2M, a generative transformer architecture designed to model the progression of human disease. Its generative capacity enables researchers to simulate synthetic future health trajectories, offering meaningful estimates of potential disease burden for up to 20 years [2]. Beyond clinical prediction, LLMs have demonstrated rapid progress in generating structured and often executable code in widely used programming languages. Models such as ChatGPT (OpenAI), Claude (Anthropic), Gemini (Google/DeepMind), DeepSeek, and others are now being applied to a broad spectrum of tasks, including the automation of data analysis. Their ability to interpret natural language instructions, process tabular data, and generate valid code has the potential to fundamentally transform how analysts and researchers interact with structured biomedical data.

This potential is especially relevant in medical informatics and public health, where there is often a need to quickly generate basic overviews and analyses from large, standardized structured datasets. In the Czech Republic, the National Health Information Portal (NZIP) provides regular openly-available datasets [3], including aggregated summaries such as age, sex, region and ICD-10 diagnoses (10th revision of the International Classification of Diseases). A methodological guide accompanies this data and includes examples of typical analytical queries (e.g., age-sex distribution of diagnoses, year-over-year comparisons of incidence etc.) [4].

Public health agencies increasingly make aggregated outputs derived from primary healthcare data openly available, in line with global trends toward data transparency and open-access public reporting. Although these datasets represent secondary analyses rather than raw patient-level records, they remain a valuable source of information for exploring epidemiological patterns, associations, and even more complex relational structures. Yet extracting such insights typically requires familiarity with dataset schemas, analytical reasoning, and practical skills in SQL or Python to formulate correct queries and transformations. As the volume of these datasets grows, non-expert users may struggle to navigate their structure or to recognize the analytical opportunities they provide, often limiting themselves to simple univariate or within-table summaries. This raises an important question: can modern large language models move beyond basic interpretation of single-table aggregates and assist in identifying, linking, and analysing more complex associations across multiple public health data sources?

A growing body of literature highlights increasing interest in the application of LLMs for similar or related questions. Galimzyanov et al. introduced PandasPlotBench [5], a benchmark designed to evaluate the ability of LLMs to generate visualizations directly from Pandas DataFrames. Their findings demonstrated that models such as GPT-4 and Claude 3.5 can automatically produce functional code for both data analysis and visualization tasks. Similarly, the LLM4DS study [6] conducted a controlled experiment with four AI assistants (Copilot/GPT-4 Turbo, ChatGPT, Claude 3.5, and Perplexity Lab) across 100 data science tasks. The evaluation measured success rates of generated code, execution validity, correctness of outputs, and the extent of manual correction required. While ChatGPT and Claude outperformed a 60% baseline, none of the tested models consistently exceeded 70% accuracy. In the biomedical domain, LLMs are being explored as tools for exploratory data analysis. A recent study reported that ChatGPT could assist researchers with limited programming expertise in conducting medical statistics analyses [7]. However, the authors emphasized the need for careful prompt design and human oversight to ensure validity and reproducibility.

Another line of research focuses on text-to-SQL translation, where natural language queries are converted into executable SQL commands, thereby facilitating database access for non-technical users [8]. This approach has also been adapted for healthcare. One study developed a graph-empowered text-to-SQL framework for electronic medical records [9], while another proposed a transformer-based model for healthcare-specific text-to-SQL conversion [10]. Complementing these efforts, [11] introduced an LLM-driven framework aimed at democratizing database access more broadly for non-technical users.

We conducted a comparative evaluation of 11 widely used LLMs on the task of generating Python code (using the pandas library) to answer analytical queries over NZIP CSV datasets. Building on prior work in program synthesis [12] and automated query generation from tabular data [13], our study assesses each model’s ability to produce correct, executable code that yields results consistent with reference outputs, with minimal or no human intervention. There are three objectives under this goal.

### Objective 1: “Ground truth replication.”

Models are required to reproduce analytical queries exactly as defined in the NZIP methodological documentation.

### Objective 2: “Go beyond replication.”

Since LLMs may have been exposed to documentation or related materials during training, we assess whether models can propose modified or generalized query formulations that remain valid and provide complementary insights beyond the reference queries.

### Objective 3: “End-to-end assistance.”

In scenarios where models have no prior knowledge of the dataset, we evaluate their ability to act as an end-to-end assistant. That is, to understand a user’s natural language question, identify appropriate dataset and relevant data columns, and generate code that executes correctly and returns meaningful results.

## 2. Methods

### 2.1. National Health Information Portal (NZIP)

The NZIP currently comprises more than 100 open datasets (as of Q4 2025), providing aggregated and anonymized information on various aspects of healthcare delivery in Czechia. For the purposes of this study, we primarily focused on the dataset Overview of Reported Diagnoses by ICD-10 Subchapters [4], as it includes a set of well-documented example analytical queries available directly on the dataset’s website.

For Objective 1, we selected four representative analytical tasks based on the methodology described in the official documentation [14]. These included, for example, calculating the number of patients with a specific diagnosis stratified by age, and assessing year-over-year differences in case counts. For each task, we considered a reference scenario solution against which LLM-generated code could be evaluated. To simulate realistic usage, all analytical queries were formulated in natural language, reflecting how healthcare professionals or analysts typically articulate questions when exploring population-level datasets. Details of the extended tasks for Objective 2 and the more exploratory use cases under Objective 3 are provided in the corresponding sections. Importantly, for Objectives 1 and 2, the language models were evaluated without access to the Internet, ensuring that they relied solely on their pre-trained knowledge and internal reasoning capabilities. In contrast, Objective 3 required Internet-enabled access, as the models needed to identify potentially relevant external datasets and supplementary information to address broader research questions.

The reference solutions were informed by our experience working with Czech healthcare data and were designed through a consensus among the contributing data scientists (GD, RS, JZ, OK).

#### 2.1.1. Analytical Queries for Objective 1: “Ground truth”

Extracted and reformulated from [4, 14].

1. How many individuals were diagnosed with Diabetes mellitus (ICD-10: E10–E14) in 2019?
2. What was the average age of individuals diagnosed with Diabetes mellitus (ICD-10: E10–E14) in 2019?
3. Is the number of individuals diagnosed with Diabetes mellitus (ICD-10: E10–E14) increasing steadily since 2010?
4. In which age groups was the lowest and highest rate of Diabetes mellitus (ICD-10: E10–E14) diagnoses per patient?

#### 2.1.2. Analytical Queries for Objective 2: “Go beyond”

In addition to strictly reproducing documented queries, we also explored modified analytical questions inspired by, but not directly formulated in, the NZIP methodological documentation [4, 14]. These tasks assess whether LLMs can generalize beyond predefined examples and generate valid code for related, but novel, queries. Such queries test the models’ capacity not only to replicate reference analyses but also to extend them to clinically relevant variations, reflecting realistic use cases where researchers may seek insights that go beyond official documentation. We examined questions such as:

1. How many individuals in 2022 had the subchapter Ischemic Heart Disease (I20–I25) recorded as a primary diagnosis, and how many as a secondary diagnosis?
2. What was the difference in mean age and regional distribution among these patients?

#### 2.1.3. Analytical Query for Objective 3: “End-to-end”

In this case, we did not predefine a specific dataset. Instead, the analytical question was intentionally formulated in a more general manner, serving as a use case for LLMs acting as research assistants rather than strict code generators. The guiding hypothesis was that two events reflected in the data may have influenced the early detection of lung cancer at stage I.:

1. An increased number of stage I. lung cancer cases diagnosed during the COVID-19 period.
2. A further increase in stage I. detections following the implementation of a national lung cancer screening program.

Based on this premise, we asked the models to consider the following questions:

1. Which datasets could be used to address these hypotheses?
2. What are the potential limitations of such data?
3. Can any of these questions be directly answered through executable Python code?

This formulation reflected a realistic research workflow in which analysts may not have complete prior knowledge of available datasets. Instead, they rely on LLMs for guidance in data discovery, methodological framing, and initial computational steps, testing the models’ ability to function as end-to-end research assistants.

### 2.2. Large Language Models

We evaluated 11 state-of-the-art LLMs across four major providers (Table 1): OpenAI, Anthropic, Google, and DeepSeek.

**Table 1.** Overview of evaluated large language models.

| Model Name | Version / Release Date | Parameter Count<br>(approx.; community estimates) | Strengths and Limitations |
| --- | --- | --- | --- |
| OpenAI o3 | 2025-04-16 | Not disclosed (≤ GPT-4) | Fast responses, high factual accuracy, suitable for structured tasks. May underperform on complex reasoning compared to GPT-4o. |
| GPT-4o | 2024-11-20 | Not disclosed | Multimodal, highly balanced performance across reasoning, factuality, and language understanding. |
| GPT-4.1 mini | 2025-04-14 | Not disclosed (smallest GPT-4 variant) | Lightweight, efficient, performs well on simple structured tasks, but lower robustness in ambiguous queries. |
| **Claude 3.5 Sonnet** | V2 (early 2025) | ~70B** | Strong reasoning and summarization, but slightly verbose. |
| **Claude 3.7 Sonnet** | 2025-02-19 | ~70B** | Improved speed and factual precision over 3.5, with strong task consistency. |
| **Claude 4 Sonnet** | 2025-05 | Estimated 100B+** | Top-tier performance in long-context understanding, precise analytical outputs. |
| **Gemini 1.5 Flash** | Version 002 | ~30–60B (est.)** | Fastest Gemini model, cost-efficient, less consistent on complex reasoning tasks. |
| **Gemini 1.5 Pro** | Version 002 | Estimated 100–130B** | Balanced across reasoning, language generation, and data tasks. Supports grounding via search. |
| **Gemini 2.5 Flash** | Preview 2025-05-20 | ~30–60B (est.)** | Faster than previous Flash version, improved factuality. Still optimized for speed over depth. |
| **Gemini 2.5 Pro** | Preview 2025-05-06 | Estimated 100–130B** | State-of-the-art factuality, strong performance in structured queries. Best suited for high-precision tasks. |
| **DeepSeek-R1** | 2025 | 671B | DeepSeek-R1 uses a Mixture of Experts (MoE) architecture. High performance on code and tabular reasoning. Slightly weaker on conversational fluency. Open-sourced and could be self-hosted. |

*\*\*neither Anthropic nor Google has officially disclosed these figures*

#### 2.2.1. Prompt Engineering

For Objectives 1 and 2, all models were provided with the particular natural language prompt, formulated primarily in Czech (full English translations are available in the Supplementary Materials). To ensure consistency and minimize stochastic variation, all queries were submitted with a fixed temperature setting of 0 (with the exception of OpenAI’s o3 model, for which this parameter was not applicable). In addition to the query text, each model was supplied with the official dataset documentation in PDF format, which describes the dataset structure and variable definitions. At the end of each prompt, we appended a few-shot example consisting of the first five rows of the dataset in CSV format. This snippet illustrated the column structure and data types, providing contextual grounding to support the model’s interpretation of the dataset and to approximate realistic analytical conditions. We did not separately define a system prompt (e.g., “You are a skilled biostatistician…”). Instead, we relied on the task formulation and contextual information to guide the model’s behavior. For Objective 3, a different prompting strategy was employed, reflecting the end-to-end assistant use case. The full prompt text for this objective is also documented in the Supplementary Materials.

#### 2.2.2. Script Execution

All generated scripts were executed locally on a MacBook Air equipped with an Apple M1 processor and 8 GB RAM, using a standard local installation of Python. For Objectives 1 and 2, the execution environment was identical across models to ensure comparability. Each script was run against the dataset nrhzs-mkn-podkapitoly.csv (file size: 223 MB). This setup allowed us to assess not only the syntactic validity of the generated code but also its practical executability on a mid-range consumer device, representative of what many analysts in public health institutions may have available.

#### 2.2.3. Evaluation Criteria

Model performance was assessed using a combination of technical metrics and domain-specific criteria, reflecting both general code reliability and the ability to produce clinically meaningful analytical results. Linguistic aspects such as code style, formatting, or inclusion of comments were not part of the evaluation, as the focus was strictly on analytical accuracy and technical reliability.

For Objectives 1 and 2, three core evaluation metrics were defined:

● Executability – Whether the generated Python code could be executed as-is without modification. We recorded whether the code:

○ ran successfully on the first attempt,
○ required minor clarification or prompt adjustment, or
○ failed completely after multiple retries.
● Correctness of results – Whether the model’s output matched the predefined reference scenario. This was assessed using binary scoring (correct/incorrect) and validated against expected values.
● Need for human intervention – Whether manual correction or clarification was required to obtain a functioning and accurate solution.

In addition, each model output was evaluated against the following specific criteria:

● Syntactic validity: The code must conform to Python syntax and be free of execution-blocking errors (e.g., SyntaxError, KeyError).
● Logical correctness: The code must implement the intended analytical logic. This included correct filtering (e.g., by ICD-10 subchapter E10–E14), accurate aggregation (e.g., by age group), and appropriate computation (e.g., use of weighted averages for age).
● Task completion: For each analytical query, we recorded whether the model attempted an answer (yes/no), and if so, whether the output was accurate.
● Use of supplementary information: We evaluated whether the model correctly incorporated details from the dataset documentation (PDF), such as distinguishing between primary and secondary diagnoses or using the correct ICD-10 codes.

For Objective 3, evaluation focused on the end-to-end assistant use case, where outputs were more explanatory and descriptive rather than purely computational. Here, model responses were judged on relevance, correctness, and coherence.

## 3. Results

The generated scripts used in this evaluation are publicly available at [15], for a detailed inspection. The repository may additionally be found via a web search using the article DOI and the keyword “git”.

### 3.1. Objective 1: “Ground truth”

The performance of all 11 LLMs was evaluated across four analytical queries, using the criteria defined in Section 2.2.3. For all models, execution time remained below one minute per task, indicating that computational efficiency was not a limiting factor. However, substantial variation was observed in the accuracy and reliability of the generated outputs.

ChatGPT-4o achieved the highest overall success rate, producing correct outputs consistently across tasks. Its performance was closely followed by GPT-4.1 mini. Both models were able to generate syntactically valid and executable code, while exhibiting robust analytical reasoning. They performed particularly well on queries involving computation of weighted averages, accurate filtering by ICD-10 diagnosis codes, and correct identification of primary diagnosis type. In contrast, the Claude models exhibited lower consistency. They frequently encountered difficulties with precise filtering for primary diagnoses, leading to incorrect or incomplete outputs. Other models displayed varying levels of robustness. While some were able to generalize effectively from the PDF documentation and the few-shot CSV examples, others required manual intervention to correct logical or syntactic errors. The OpenAI o3 model was a notable outlier, failing to execute successfully despite repeated attempts and prompt restarts.

A comprehensive comparison of model performance, including executability, result accuracy, and task-specific capabilities, is presented in Table 2.

**Table 2.**
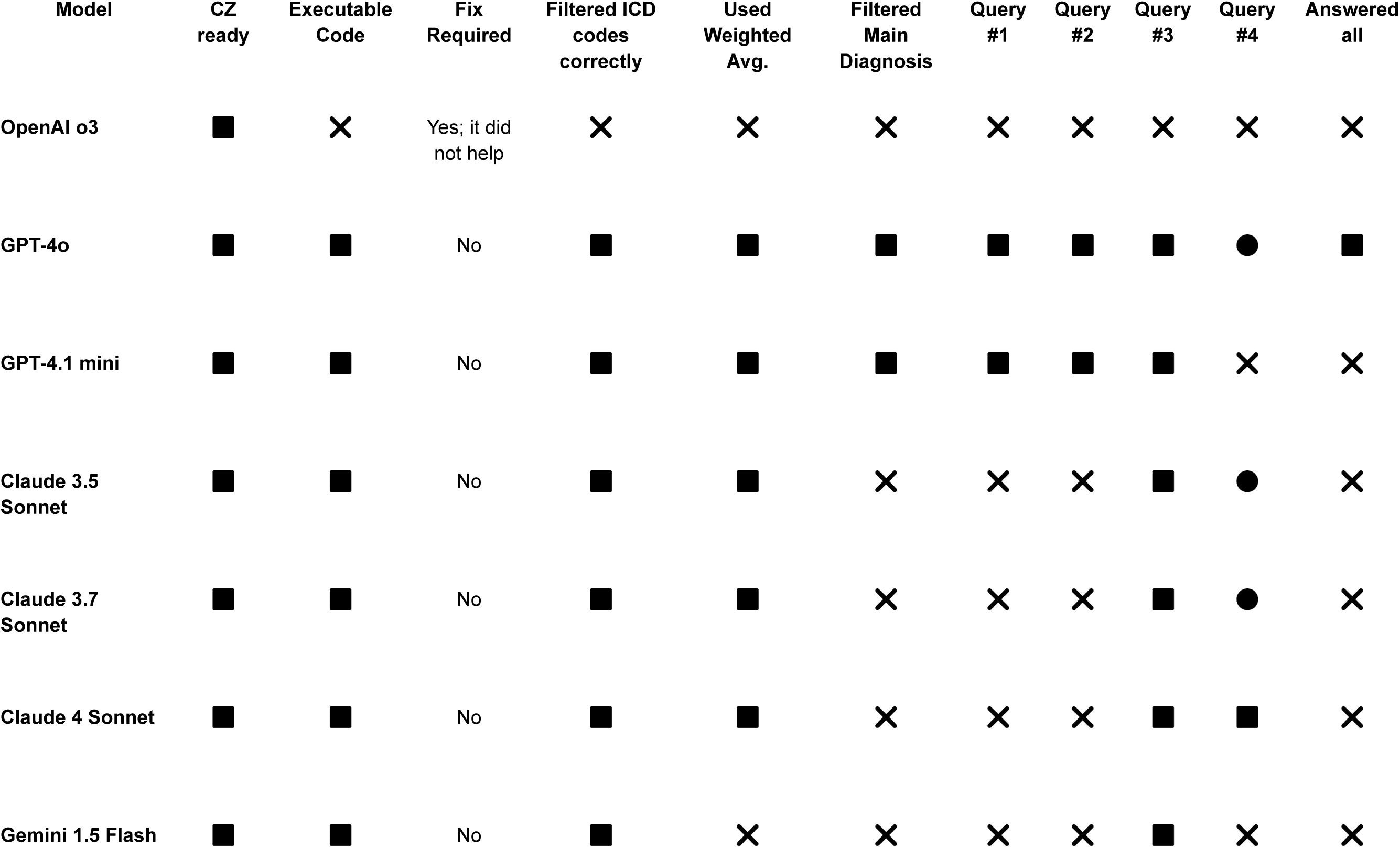
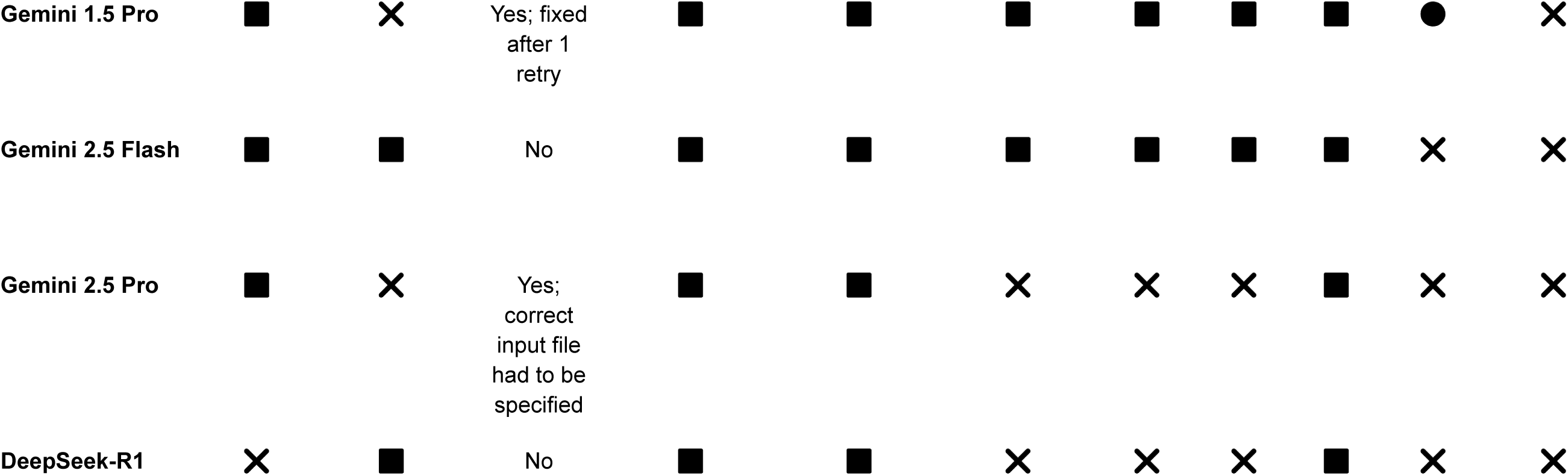
Objective 1: Model performance across analytical tasks. ✅ = Fully met, ❌ = Not met, •= Partially met.

#### 3.1.1. Caveats

During the evaluation, sub-question #4 emerged as potentially problematic. It was not entirely clear what the “correct” interpretation should be. Several models directly reported the categories exactly as they appeared in the provided dataset. In such cases, we marked the answer as Partially met rather than Fully met, because this approach did not fully satisfy the intended requirement. The core issue lies in the discrepancy between age categories as given in the dataset and the actual age derived from birth years.

Notably, the instruction “Split the data by age categories (e.g., 10–14 years, 95+ years) and report the results” was correctly followed only by Claude 4 Sonnet, as far as we observed. Ideally, in our opinion, the data should be grouped by the age in years, not categories; otherwise, each patient could be counted multiple times (in our example, up to ten times). Beyond this, the task leaves room for interpretation. For instance, the oldest age category naturally contains the smallest number of patients in absolute terms but represents the highest share relative to its underlying population segment.

Taken together, these issues indicate that the question itself should be more precisely defined and this highlights an important finding, i.e. when a task formulated in natural language is ambiguous or incomplete, it may be difficult to determine a single fully correct solution. On the other hand, the solutions suggested by LLMs can help reveal ambiguities or flaws in the task definition that might otherwise remain unnoticed.

### 3.2. Objective 2: “Go beyond”

In contrast to Objective 1, where OpenAI o3 failed completely, it paradoxically achieved the best performance in Objective 2, successfully meeting all evaluation criteria. Across the other models, the most frequent source of error was the incorrect handling of diagnosis type (main vs. secondary). Only o3 consistently separated diagnoses as required. GPT-4o, GPT-4.1 mini, and the Claude family generally executed code correctly but often failed to implement this distinction, leading to partial rather than complete solutions. Gemini 2.5 Pro failed execution entirely due to an ImportError (“Missing optional dependency ‘Jinja2’”), which arises from the .style method in Pandas. Since Jinja2 is not a standard library dependency and cannot be assumed available in most baseline Python installations, we did not count this case as a valid solution. A difference compared to Objective 1 was observed with DeepSeek-R1, which in this case interacted in Czech.

Overall, for basic executability, most models performed well. A comprehensive comparison of model performance, including executability, result accuracy, and task-specific capabilities, is presented in Table 3.

**Table 3.**
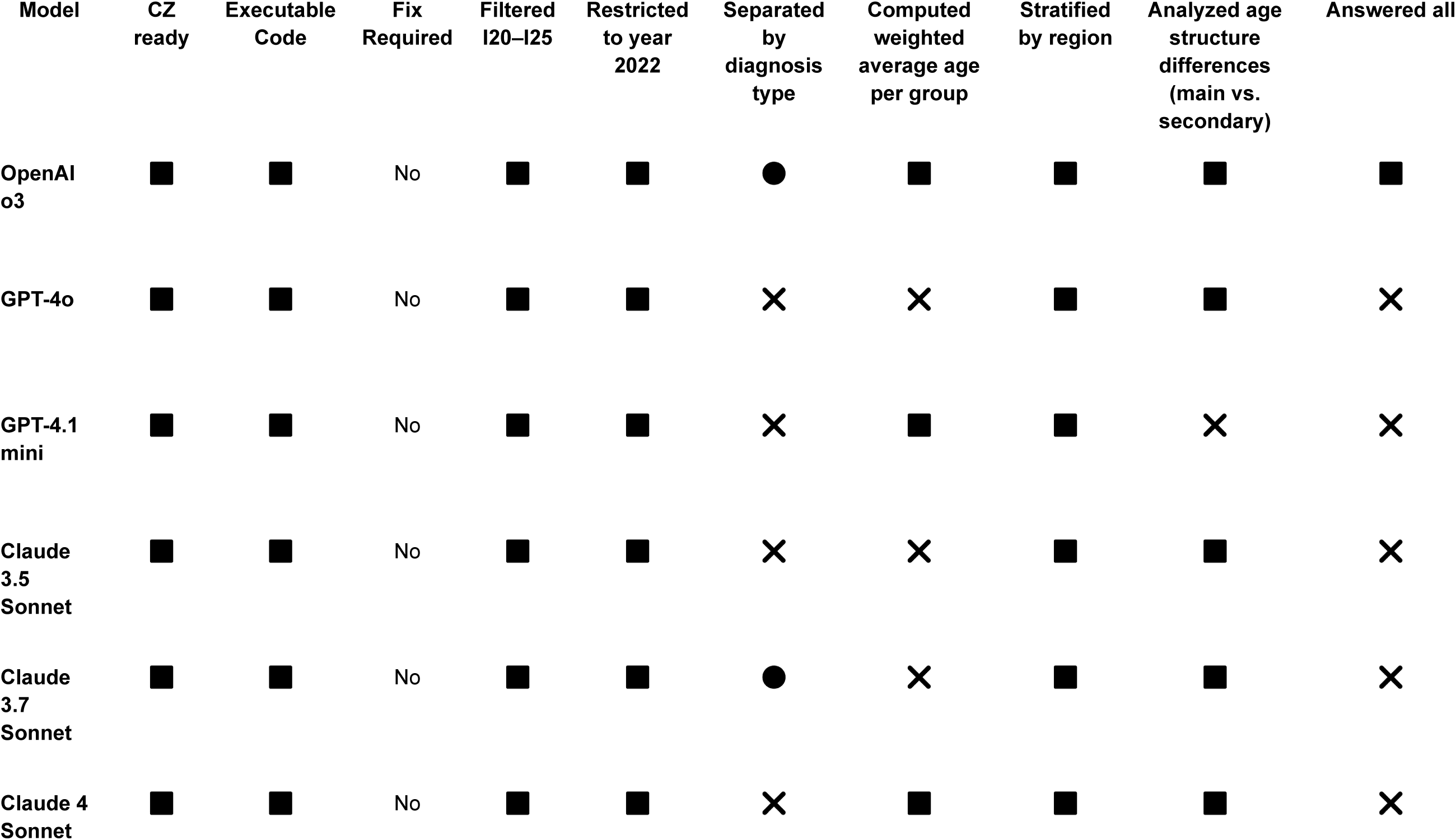
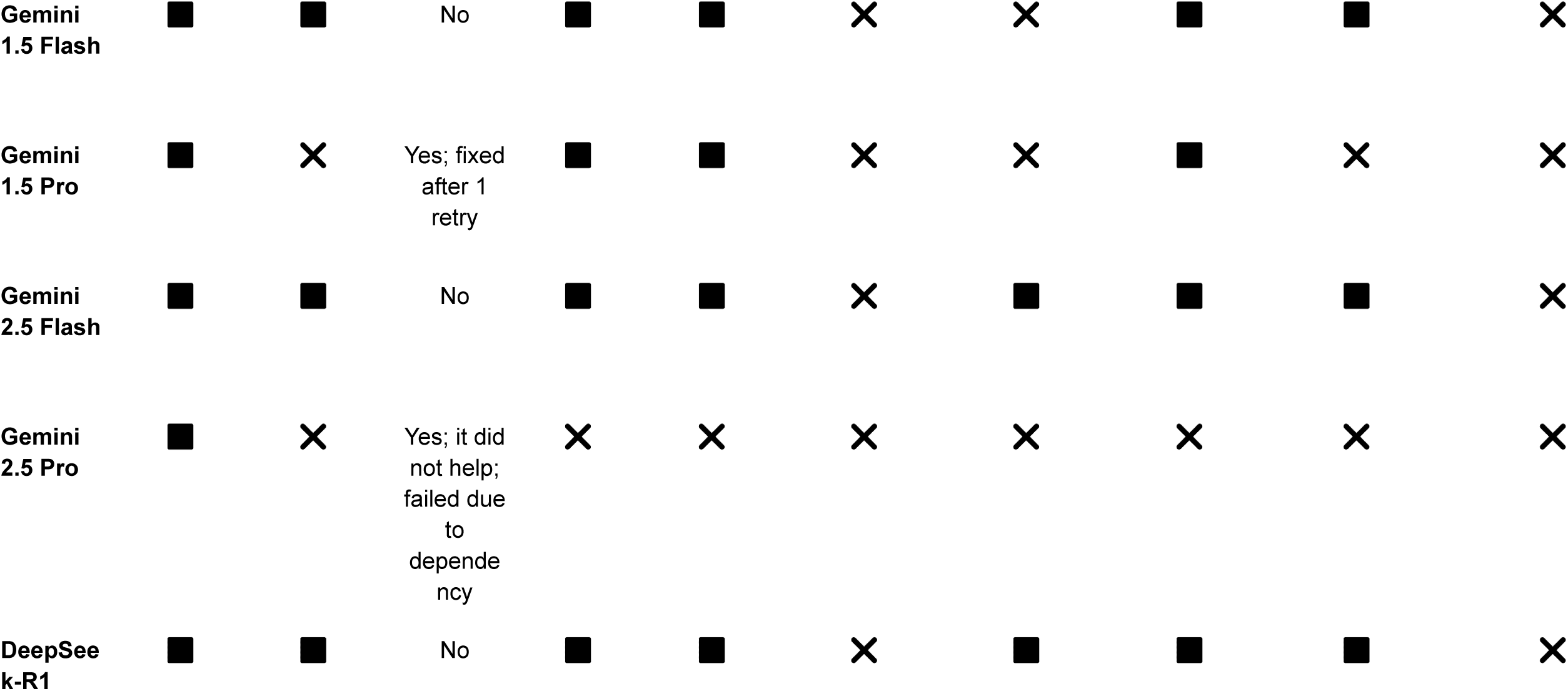
Objective 2: Model performance across analytical tasks. ✅ = Fully met, ❌ = Not met, •= Partially met.

#### 3.2.1. Caveats

The sub-question “Separated by diagnosis type” proved non-trivial to evaluate. It is clear, and represents a strength of several of the evaluated models, that they correctly interpreted the documentation distinguishing primary (ZDG) and secondary (DG) diagnoses. However, the underlying database ultimately does not permit a direct answer to the question as it was originally formulated. For this reason, we marked the corresponding solutions as Partially met.

### 3.3. Objective 3: “End-to-end”

For Objective 3, we considered only one model, ChatGPT-4o. This model was selected because it ranked among the best performers in Objective 1 and, importantly, it was the only one in our study with Internet access (via Microsoft Copilot). The first step was to test whether the LLM could, when provided with the root directory URL of the NZIP datasets, identify the correct dataset on cancer incidence, or otherwise recommend relevant datasets. The model successfully returned three relevant datasets (Fig. 1).

**Fig. 1.**
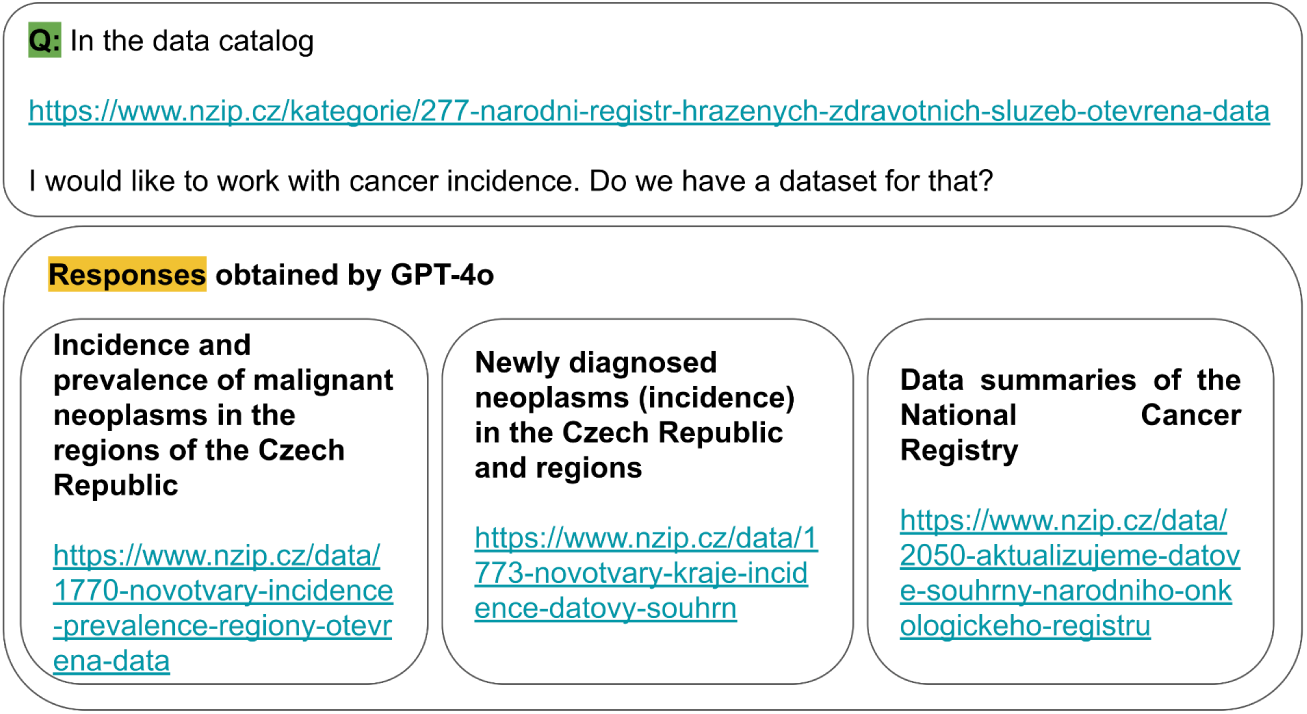
Example of Objective 3 input prompt and LLM output (dataset identification).

A second prompt, containing concrete research questions, was then submitted. The full text of this exchange is provided in Fig. 2. In this case, the model not only identified suitable datasets but also generated a limitations section (as explicitly required in the prompt), which included points such as:

● Data lag: The most recent data may only be available up to 2022 or 2023, limiting long-term evaluation of screening impact.
● Missing variables: The dataset does not explicitly indicate whether a cancer case was detected through screening or incidentally.
● COVID effect: The increased number of stage I lung cancer detections during the COVID-19 pandemic may be confounded by other factors, such as more frequent chest CT scans.
● Aggregation: Some datasets are aggregated, which constrains detailed statistical analyses.

**Fig. 2.**
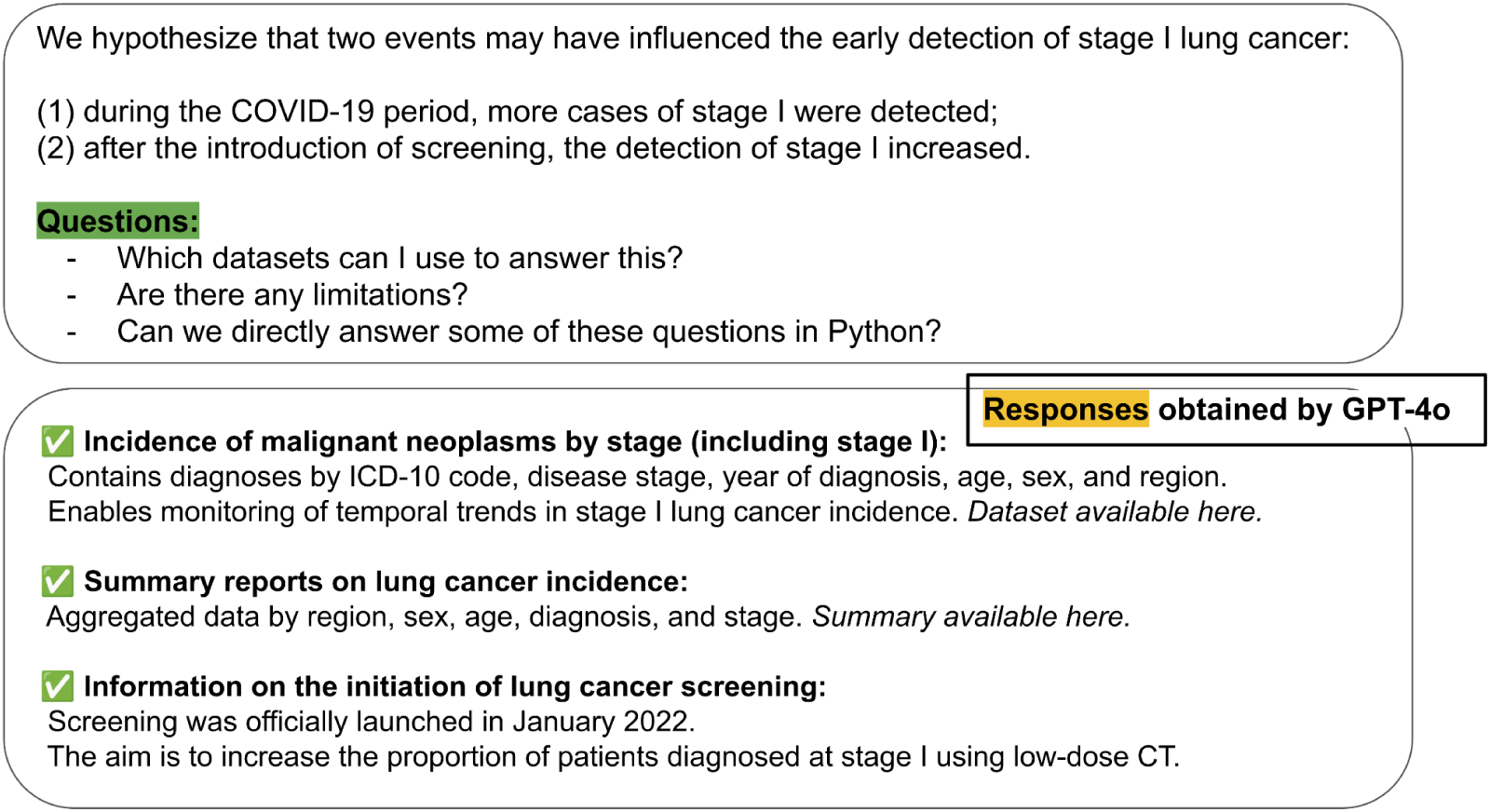
Objective 3 Input–Output example: Stage I lung cancer incidence.

In addition to descriptive text, the model generated executable Python code. The script implemented the following analytical tasks:

● Analysis of trends in stage I lung cancer incidence over time (2015–2023).
● Comparison across three predefined time periods:

○ Pre-COVID (2015–2019),
○ COVID period (2020–2021),
○ Post-screening introduction (2022–2023).
● Stratification of cases by region (NUTS level) and age groups (10-year intervals).
● Suggested calculation of absolute case counts and incidence rates per 100,000 inhabitants.

The generated code relied only on columns that exist in the open NZIP datasets: *diagnoza_kod, stadium, rok_dg, kraj_nazev, vek*. Overall, it correctly implemented filtering for diagnosis code C34 (lung cancer) and stage I, and then computed case counts by region and age group. These counts were suggested to be merged with external population data from the Czech Statistical Office (ČSÚ) in order to calculate incidence rates. Finally, the script generated a heatmap visualization and exported the aggregated results to CSV format.

#### 3.3.1. Strengths and Caveats

From a qualitative perspective, the model successfully performed the three intended roles for an end-to-end assistant, i.e. dataset discovery, critical appraisal (limitations), and generation of an implementable analysis plan and script. The script explicitly used only the columns present in the NZIP data and included references to external population data sources needed to compute incidence rates. However, several important caveats must be emphasized for the obtained results:

● Although the LLM returned plausible dataset names and links, provenance and exact file contents must be validated by a human researcher before executing any analysis. LLMs can hallucinate filenames or attribute semantics incorrectly even when Internet-enabled.
● As the model itself noted, the latest available NZIP data may lag (e.g., through 2022–2023). Any inference about screening effects or COVID impacts therefore requires careful consideration of data completeness and temporal alignment.
● The NZIP data do not contain an explicit variable indicating whether a diagnosis resulted from screening; the model correctly flagged this as a limitation. Consequently, causal claims about the effect of screening cannot be made from these data alone without linkage to screening registries or patient-level screening variables.
● Regional mapping (NUTS mapping) and population denominators require careful reconciliation (e.g. consistent region codes and age-group definitions) to avoid misestimation of incidence rates.

## 4. Discussion

In this study, we demonstrated that large language models (LLMs) are capable of generating executable, often fully functional Python code for data analysis tasks relevant to aggregated health datasets provided to the public. In many cases, the models produced correct outputs without requiring manual modification, successfully interpreting the CSV schema and applying appropriate analytical logic.

One of the most important lessons learned for Objective 1 is that state-of-the-art models such as ChatGPT-4o and GPT-4.1 mini handled typical analytical tasks (e.g., filtering, aggregation, calculating weighted averages) with high reliability, even in a non-English context. The presence of diacritics, local column names, and Czech-specific medical terminology posed no substantial barrier. Most models processed the few-shot examples correctly and adapted to the structure of the data with remarkable accuracy. A surprising and encouraging finding was that the majority of outputs were not only syntactically valid but also executable without errors. This suggests that LLMs may already be suitable as assistants in the early stages of analytical pipeline development, particularly in contexts where users need support in transforming natural-language questions into executable code. Indirectly consistent with our findings, a recent evaluation on Brazilian medical aptitude exams (non-English context) similarly found that GPT-4o achieved the best performance, and in addition to the observed results herein, large-scale proprietary models substantially outperformed open-source models such as Qwen and LLaMA [16]. In another 2025 study [17], the authors showed that when using carefully designed prompt-engineering strategies, GPT-4o ranked among the top performers in validating drug-repurposing hypotheses.

Nevertheless, performance varied across models. Claude and Gemini, for example, frequently failed to apply the required filtering for primary diagnoses, despite this instruction being explicitly stated in the input prompt. This limitation illustrates that even when models can parse code structure, they may overlook essential logic embedded in the task description. Interestingly, Gemini 1.5 Pro was able to correct such mistakes in subsequent queries, suggesting potential for intra-session adaptation. DeepSeek-R1, in contrast, consistently generated responses in English rather than Czech, which may limit its utility in public sector or healthcare contexts where local language use is essential. It is important to note that, although DeepSeek-R1 is currently prohibited for production use under many institution policies, we still considered it essential to include the model in our benchmarking because it is one of the few high-performing LLMs that can be fully self-hosted. Nevertheless, real-world adoption in our context should prioritize models that combine self-hosting capability with robust local-language support and compliance with institutional security requirements (e.g., secure variants of LLaMA, Qwen, or other models validated for privacy-preserving deployment).

Some tasks further revealed that models could generate syntactically valid and executable code that nonetheless failed to match the expected results. This was particularly evident in Query 1 and Query 2, where the outputs “looked correct” but returned erroneous answers. These subtler forms of failure highlight the necessity of systematic validation processes, even when the generated code executes without error.

For Objective 2 (query customization), we observed a moderate degree of variability across models. In practice, it is often necessary to explicitly instruct an LLM to perform certain operations, such as calculating a weighted average, if this is not done automatically. Encouragingly, providing clarifications in Czech often resulted in immediate corrections, demonstrating that models can adapt rapidly within a single session. For non-programmer analysts, this type of interaction is not a significant barrier, but rather a natural part of iterative query refinement.

For Objective 3, we explored a more open-ended, end-to-end use case, where the model was not provided with a predefined dataset but instead had to identify suitable resources, acknowledge their limitations, and propose an analytical strategy. ChatGPT-4o demonstrated the ability to navigate the NZIP catalog, retrieve relevant datasets, and generate both descriptive limitations and executable Python code tailored to the research question. The returned outputs were not technically incorrect but were contextually relevant, incorporating epidemiological reasoning such as COVID-related diagnostic effects, screening program initiation, and the constraints of aggregated data.

To our knowledge, this is the first systematic comparison of LLMs applied to real-world public health queries over structured healthcare datasets. Related work has evaluated LLMs in adjacent areas, such as their performance in medical education. For example, a recent comparison of DeepSeek-R1 and ChatGPT-4o on the Chinese National Medical Licensing Examination [18] illustrates the growing interest in benchmarking LLMs within healthcare-specific contexts.

An important methodological question concerns the reproducibility of results across repeated runs. Since ChatGPT-4o emerged as one of the best-performing models in our study, we examined whether its outputs remained stable when identical prompts were re-issued. A previous study [19] reported that while the free version of ChatGPT showed variability over time, the paid versions remained stable, with no significant changes across updates. Consistent with these findings, another study on information extraction from electronic health records (EHR) [20] found that GPT-4o produced perfectly reproducible outputs across repeated runs at temperature 0. In our own non-systematic, selectively chosen retest attempts, issuing the same prompt from scratch generally produced nearly identical code and the same numerical outputs, supporting the view that averaging over multiple runs may be unnecessary for this class of analytical tasks. However, broader testing would be needed to generalize this observation. Since temperature=0 was used, results should be deterministic.

A key limitation of this study is the scope and complexity of the analytical queries. We focused primarily on well-defined, moderately complex tasks over a single class of public health datasets. Future work should expand the set of tasks to include more advanced analytical operations, such as multi-step aggregations, time-series modeling, and predictive analyses, in order to better assess the upper limits of LLM capabilities in applied public health informatics. While execution time was uniformly fast (under one minute per query), our study did not systematically evaluate performance trade-offs under conditions involving larger datasets, high-dimensional tables, or real-time constraints. Several recent studies highlight the potential for resource-efficient LLMs in data analysis. For example, López Espejel et al. (2024) demonstrated that smaller, open-source models can achieve competitive performance when properly configured [21]. Their findings suggest that lower-cost models could be leveraged for public health analyses, particularly when local deployment or privacy considerations preclude the use of commercial cloud models.

Another important consideration is knowledge verification. Generative LLM outputs, while syntactically and logically plausible, must be validated before adoption in clinical or epidemiological analyses. Recent work in biomedical generative AI [22] demonstrates that models can generate disease-centric associations and structured biomedical knowledge, but ontologies and curated vocabularies are essential to verify terms for diseases, drugs, symptoms, and genetic relationships. Translating this principle to public health data analysis implies that generated code and aggregated results should be checked against official coding standards (e.g. ICD-10), population registries, or other authoritative references.

Related software solutions in the EHR domain illustrated practical approaches to end-to-end reasoning and query generation. For example, ClinQuery [23] implemented a pipeline from named entity recognition to SQL query generation and natural language reasoning, enabling retrieval of insights from complex clinical datasets. Similarly, graphRagSqlator [24] can explore database schemas automatically and generate valid SQL queries from natural language prompts. These systems are designed to handle large databases with heterogeneous table structures, i.e. possibly challenges analogous to those faced when working with NZIP or other multi-table public health registries. By contrast, our work demonstrates that LLMs can already generate fully functional Python code over structured CSV datasets, providing a foundation for future extension to larger, relational patient-level datasets.

Our findings suggest that contemporary LLMs are increasingly capable not only of generating technically correct analytical code, but also of interpreting the clinical meaning of aggregated health indicators and identifying variables that are relevant to complex associative questions. Remarkably, several models demonstrated an ability to reformulate natural language requests into multi-step analytical strategies that approximate human reasoning about disease patterns, diagnostic pathways, or temporal dynamics. Even when working with aggregated data, where clinical nuance is inherently constrained, the models were often able to infer which dimensions (e.g., age, sex, diagnosis type, region, or temporal trends) were likely to carry meaningful epidemiological information. This capability has profound implications. As public health agencies continue to expand the availability of open aggregated datasets, LLMs could substantially empower citizens, journalists, clinicians, and policymakers who lack formal training in analytics, effectively lowering the threshold for accessing and interpreting population health trends. At an institutional level, these technologies raise the possibility of shifting human analysts away from manual coding and routine exploratory tasks toward higher-value work such as designing richer and more semantically coherent datasets, curating metadata, and developing synthetic datasets that mirror patient-level structures such as disease history. In such a future ecosystem, LLMs may function as scalable analytical engines, while human experts focus on ensuring data integrity, ontology alignment, and interpretability. Ultimately enabling more sophisticated, reproducible, and clinically meaningful analyses across the public health data landscape.

## 5. Conclusion

This study demonstrates that general-purpose LLMs can serve as effective coding assistants for public health data analysis, including tasks involving Czech-language input and local healthcare data structures. Even the most capable models, however, require post hoc verification and cannot yet be relied upon without human oversight.

The concept of “LLM-as-a-coder” for public health analysts appears promising, particularly when combined with validation mechanisms and carefully designed prompts. With increasing reliability, multi-language support, and fast inference times, these tools have the potential to accelerate the development of analytical pipelines and reduce the technical barrier for non-programmer analysts.

Looking forward, we recommend further exploration of hybrid human–AI workflows, in which analysts use LLMs to bootstrap code that is subsequently refined via automated testing and expert review. Future studies should also assess integration with structured health data platforms, such as OMOP Common Data Model (CDM) or NZIP systems (Syntetika), and investigate models’ adaptability to domain-specific ontologies and coding standards. We further anticipate that retrieval-augmented generation (RAG) approaches could improve LLM performance, particularly in cases requiring access to large or dynamically updated datasets, by combining generative reasoning with verified data retrieval. Also, given the documented failures with PDF interpretation, testing RAG-enhanced approaches would be a natural follow-up.

## Data Availability

Scripts and data produced within this study are available online at https://gitlab.fel.cvut.cz/klempond/nzip-llms-eval. If applicable, any other data produced in the present study are available upon reasonable request to the authors.

## Acknowledgement

This research was supported by the project CZ.02.01.01/00/23_025/0008743, funded by the European Union under the Operational Programme Johannes Amos Comenius (OP JAK).

## Supplement Objective 1 – Prompt in English (translation)

### Task

Based on the data structure and information provided in the PDF, answer the queries below. You are given data in CSV format containing reported diagnoses by ICD-10 subchapters. Prepare Python code using the Pandas library that can be used to answer the following queries from the given data:

- Determine the trend in the number of individuals diagnosed with the subchapter Diabetes mellitus (E10–E14) from 2010 to 2019 and verify whether this number is steadily increasing. Provide specific counts of individuals for each year during this period.
- Find out what the number of individuals diagnosed with the chapter Diabetes mellitus (E10–E14) was in 2019, and determine the average age of these patients.
- Identify the age category in which the highest and the lowest number of diagnoses of Diabetes mellitus (E10–E14) per patient occurred. Divide the data into age categories (e.g., 10–14 years, 95+ years) and report the results.

### Data context

• The data is organized according to the structure described in the PDF (ICD subchapter, year, age category, number of patients, average age). • The relevant ICD subchapter for Diabetes mellitus corresponds to codes E10–E14. • Focus on the primary type of diagnosis. • The questions relate to the outputs described in the section “Example of a specific output” and include trend analysis, aggregation by year, and aggregation by age category.

### Technical requirements

- If possible, write Python code using the Pandas library or an SQL-like query that directly answers the given questions. • State any assumptions made when creating the query or algorithm.

### Objective

Your goal is to accurately answer the questions above using algorithmic analysis that is precise, consistent, and appropriate for use with the CSV dataset. Keep in mind that the outputs will reflect your model’s ability to work with data and generate analyses based on the materials provided.

### Example of the first 5 rows of the dataset (nrhzs-icd-subchapters.csv)

year age_cat gender patient_id_district pacient_id_region ICD_subchap type_dg subjects avg_age 2017 20 1 411 41 91 ZDG 14 96.35714 2016 19 2 020C 20 80 DG 42 91.43284 2021 5 2 425 42 71 ZDG 67 21.77215 2013 8 2 635 63 40 ZDG 35 37.61111 2012 9 1 010A 10 80 DG 524 41.81461

## Supplement Objective 2 – Prompt in English (translation)

Based on the data structure and the information provided in the PDF, process the queries below. Data is available in CSV format containing reported diagnoses according to ICD-10 subchapters. Prepare Python code using the Pandas library that can generate answers to the following queries:

### Example

Main vs. secondary diagnoses in Ischemic Heart Disease

How many people in 2022 had the subchapter Ischemic Heart Disease (I20–I25) recorded as the main diagnosis and how many as the secondary?

What was the difference in average age and regional distribution?

### Evaluation subtasks

Filter diagnoses I20–I25. Narrow down to year 2022. Split by diagnosis type: main vs. secondary. Count the number of unique patients in each group. Calculate the average age for each group. Further break down by regions and create a comparison table. Determine whether the typical age structure of patients differs between main and secondary diagnoses.

### Technical requirements

• If possible, write Python code using Pandas functionality or an SQL-like query that directly answers the questions.

### Goal

Your goal is to precisely answer the queries using algorithmic analysis that is accurate, consistent, and suitable for use with the dataset in CSV format. Keep in mind that the outputs will reflect your model’s ability to work with data and generate analyses based on the provided materials.

## Supplement Objective 3 – Prompt in English (translation)

We have a hypothesis that two events may have influenced the early detection of lung cancer in stage 1. During the COVID period, more cases of lung cancer were detected in stage 1. Secondly, more cases of stage 1 were detected after the introduction of screening.

1. What datasets (or combinations of datasets) can I use to answer this?
2. Are there any limitations?
3. Provide source code in Python.

